# Effect of Rhythmic Auditory Stimulation (RAS)^®^ with and without Melody on Parkinson’s Disease (PD) Patients with Deep Brain Stimulation (DBS): A Study Protocol

**DOI:** 10.1101/2025.05.09.25327235

**Authors:** Reina Arakawa, Kyurim Kang, Isabella Sterner, Joseph Seemiller, Lauryn Currens, Rebecca Khamishon, Yousef Salimpour, Kelly Mills, Alexander Pantelyat

**Affiliations:** Department of Neurology, Johns Hopkins University School of Medicine; Center for Music and Medicine, Johns Hopkins University School of Medicine; Ludwig-Maximilians-Universität München; Department of Neurology, Penn State College of Medicine

## Abstract

Parkinson’s Disease (PD) is characterized by motor impairments, including gait abnormalities that contribute to instability and an increased risk of falls as the disease progresses. While deep brain stimulation (DBS) of the subthalamic nucleus (STN) or the globus pallidus internus (GPi) can be highly effective for managing tremor, rigidity and limb bradykinesia, its impact on gait remains limited, highlighting the need for dedicated gait therapies. Rhythmic auditory stimulation (RAS)**^®^** has been used to improve PD gait parameters, such as cadence and velocity. This pilot study aims to investigate the neurophysiological underpinnings of two RAS^TM^ approaches: pure rhythmic RAS, which uses metronome-generated beats, and melodic RAS, which incorporates composed music along with rhythmic beats. We will record local field potentials (LFP) from the DBS devices already implanted in individuals with PD as part of their routine care, along with gait parameters, to assess neurophysiological and behavioral responses to RAS. By recording neural signals from the STN and GPi simultaneously with assessment of gait parameters, this study will examine how different RAS paradigms influence motor performance and underlying brain activity. This innovative approach will provide insights into how pure and melodic RAS paradigms modulate neural activity in basal ganglia structures, advancing our understanding of RAS effects on motor function in PD.

## Introduction

Parkinson’s Disease (PD) is a complex progressive neurodegenerative illness associated with significant morbidity and adverse impacts on quality of life. While PD pathology involves degeneration of dopaminergic neurons in the substantia nigra pars compacta that underlies motor symptoms, the disease presents with a highly heterogeneous variety of symptoms (Bloem et al., 2021) related to both dopaminergic and non-dopaminergic mechanisms. Common motor symptoms of PD include asymmetric resting tremor, rigidity, bradykinesia, and changes in posture and gait, including postural instability (Postuma et al., 2015). Non-motor symptoms in PD include hallucinations, depression, anxiety, neurogenic bladder dysfunction, sleep disorders, and anosmia (Hausdorff et al., 2007).

Therapeutic management of PD typically involves pharmacological interventions aimed at restoring normal dopaminergic tone in the basal ganglia-thalamo-cortical networks that underly movement control. In cases where pharmacotherapy fails to adequately address motor symptoms, Deep Brain Stimulation (DBS) is considered for selected patients, typically targeting the subthalamic nucleus (STN) or the globus pallidus internus (GPi) (Halli-Tierney et al., 2020; Weaver et al., 2009).

While many studies have shown that STN-DBS or GPi-DBS in PD is advantageous for tremor, rigidity, and bradykinesia (Weaver et al., 2009), fewer studies have noted clear benefits for posture and gait abnormalities (Rahimpour et al., 2021), and these symptoms represent a significant unmet need. As PD progresses, addressing gait abnormalities becomes crucial as worsening motor impairments contribute to greater instability and a higher risk of falls. Therefore, additional approaches are necessary to enhance the effect of existing treatment options.

### Rhythmic Auditory Stimulation (RAS)^®^

Rhythmic auditory stimulation (RAS)^®^ is a Neurologic Music Therapy^®^ technique that involves rhythmic and/or musical stimulation to improve different aspects of movement, including gait (Gonzalez-Hoelling et al., 2021; Mainka et al., 2018; M. H. Thaut et al., 1996; M. Thaut & Hoemberg, 2014). The standardized RAS^TM^ protocol comprises the following steps: 1) determining the individual’s baseline cadence (steps/minute); 2) adjusting the metronome rhythm to match the patient’s cadence; 3) instructing the individual to synchronize their footsteps with the beat; 4) gradually increasing the pace by 5-20% above the individual’s baseline cadence; and 5) gradually fading out the RAS (M. Thaut & Hoemberg, 2014). RAS has been shown to improve several gait parameters in PD including gait velocity and stride length, and may also potentially reduce falls (Arias & Cudeiro, 2010; Calabrò et al., 2019; Murgia et al., 2018; M. H. Thaut et al., 2019; Ye et al., 2022).

PD patients are known to exhibit deficits in internal timing regulation, leading to challenges performing automated actions that require consistent pacing, such as walking (Rodger & Craig, 2016). A prevailing hypothesis across research groups suggests that RAS may facilitate network level interaction among the auditory system, cerebellum, basal ganglia, and frontal executive areas, leading to enhanced fronto-temporal-cerebellar cortical striatal network activation (Grahn, 2009; Koshimori & Thaut, 2018). This interaction is posited to enhance intra-network connectivity, potentially compensating for basal ganglia dysfunction in PD (Bella et al., 2015; Koshimori & Thaut, 2018). By providing rhythmic sound cues with predictable intervals, RAS enables gait synchronization with an external rhythm, which may aid patients in improving movement speed and trajectory and muscle recruitment (Bella et al., 2015; Braunlich et al., 2019; Calabrò et al., 2019; M. H. Thaut et al., 2014, 2015). Furthermore, research has shown that beta modulation in the STN increases when patients receive metronome-based auditory cues while stepping in a seated position, supporting more consistent step timing. (Fischer et al., 2018).

While several studies have suggested that RAS can modulate excessive cortical activity (Koshimori & Thaut, 2018; Nishida et al., 2021; Tosserams et al., 2022), the extent to which RAS modulates subcortical activity specifically in the STN or GPi remain less well understood.

### RAS versus RAS with melody

RAS intervention involves either a musical piece with accentuated and superimposed rhythmic beats (Bella et al., 2015; Calabrò et al., 2019; Gonzalez-Hoelling et al., 2021; Mainka et al., 2018) or a metronome delivering purely beat-based stimuli (Capato et al., 2020; Murgia et al., 2018). Researchers have studied various components within these approaches. For example, Capato and colleagues (2020) investigated the effect of pure beat RAS in PD patients with and without freezing of gait (FOG). Similarly, Murgia and colleagues (2018) explored differences of artificial metronome sounds and “ecological RAS,” which utilized recorded footstep sounds. While numerous studies have examined the effects of pure beat RAS or musical RAS on gait independently, research directly comparing pure beat RAS and melodic RAS is limited. This gap poses an important limitation for a complete understanding of RAS and its optimal effects, particularly in the context of individualized clinical applications, as highlighted by Rodger et al. (2016).

To specifically evaluate the contribution of rhythm and melody in RAS, this pilot study will compare two distinct RAS paradigms: a “pure RAS” approach utilizing metronome-generated drumbeats and a “melodic RAS” approach incorporating original composed music with superimposed metronome drumbeats. Both paradigms will be assessed for their impact on behavioral outcomes (e.g., gait parameters) and neurophysiological responses (e.g., Local Field Potentials, LFP).

### Local Field Potentials (LFP) activity

LFP represent synaptic potentials aggregated across groups of neurons near the recording electrode in the DBS lead, are directly associated with transmembrane currents and provide valuable insights into correlated neural activity (Herreras, 2016). While electroencephalography (EEG) provides an overview of cortical activity, LFP offer a more localized measure of neural activity, capturing signals from subcortical tissue, such as the STN and the GPi (Bush et al., 2024; Buzsáki et al., 2012). Previous studies using LFP found that certain frequency bands are often associated with specific activated neural networks both physiologically and pathologically (Friston et al., 2015). These identified frequency bands can provide a clinically relevant biomarker of brain activity; in fact, beta frequency band activity has been successfully used to develop a recently approved closed loop DBS system, which triggers stimulation in response beta frequency band changes (Stanslaski et al., 2024; Wilkins et al., 2024).

PD patients typically show a pathologically increased oscillatory synchrony in the STN in the 8 to 35 Hz range (alpha and beta range), whose suppression via dopaminergic medication or DBS leads to symptom improvement (Kühn et al., 2006a). While the alpha range (8-12 Hz) was previously shown to correlate with the intensity of dyskinesia, it has been mostly evaluated in association with cognitive function and emotion (Yin et al., 2022). Thus, the present study will primarily focus on the motor band, specifically the beta frequency band ranging from 13 to 35 Hz.

To better understand the underlying neurophysiological effects of pure rhythmic RAS and melodic RAS, this study will leverage advanced techniques to measure LFP from the DBS device already implanted in study participants with PD for clinical purposes. By capturing these neural signals directly from targeted subcortical regions alongside gait parameter outcomes, we will compare the effects of pure rhythmic beats versus rhythmic beats with melodic patterns on gait performance and underlying brain activity.

## Materials and Methods

### Participants

We will recruit participants diagnosed with idiopathic PD according to 2015 MDS Criteria (Postuma et al., 2015) and implanted (in the STN or GPi) with the Medtronic® Percept™ PC Model B35200 neurostimulator with BrainSense™ technology (Percept™ PC) or Medtronic® Percept™ RC (rechargeable neurostimulator). Participants must have had at least 1 additional follow up after their initial programming visit for inclusion. The study procedures, risks, and data collection process will be explained to all participants and written informed consent will be collected prior to study participation. Table 1 indicates inclusion and exclusion criteria for this study.

**Table 1.** Inclusion and Exclusion Criteria.

| Inclusion | Exclusion |
| --- | --- |
| Diagnosed with idiopathic PD (Postuma et al., 2015) | Participants unable to understand verbal instructions. |
| DBS implantation with Medtronic Percept PC or RC DBS Device in bilateral STN or GPi | Participants who have previously reported significant discomfort when DBS is turned off. |
| Age 18 to 95 | Participants who, in the view of the investigators, would have severely increased parkinsonian symptoms in the DBS OFF state, potentially causing intolerable discomfort or disability. |
| Fluency in English | Participants who are non-ambulatory or exhibit Hoehn and Yahr stages 4-5 in the “on” medication state |

We plan to recruit all eligible patients from the Johns Hopkins Parkinson’s Disease and Movement Disorders Center who meet the inclusion criteria described above.

### Study design

This pilot study is a repeated measures design, where each participant is exposed to the control condition (without RAS) as well as both intervention conditions (with Pure RAS and with Melodic RAS). This within-subject design allows for direct comparison of outcomes across conditions within the same individual, enhancing our ability to detect changes attributable to the intervention. Each visit will be conducted by both a study clinician and a research team member. The study clinician will monitor and manage the Medtronic Percept^TM^ tablet, respond in case of adverse events, and assess the gait and balance items of the MDS-UPDRS III (3.9 through 3.13). The researcher will measure the cadence and velocity of the participants’ gait and keep track of the different conditions. The assessment will begin with the participant’s deep brain stimulation (DBS) randomly assigned to either the ON or OFF condition. The initial stimulation status will be known only to the study clinician, while the researcher, data analyst, and participant will remain masked. Because participants may recognize their stimulation status based on perceptible differences in symptom response between the ON and OFF states, this is considered a single-masked study. An important and novel aspect of this study design involved composing a musical piece to serve as the melodic RAS intervention. In selecting the type of music for the melodic RAS intervention, the NIH Music-Based Intervention (MBI) Toolkit offers a framework for study design guidance (Edwards et al., 2023). According to the MBI Toolkit, the following elements must be considered in music used for clinical interventions: Frequency, Tempo, Melody, Playback volume, Pitch, Timbre, and Rhythm. Additional considerations include the familiarity of the music and its personalization. As the aim of this study is to compare the effects of RAS with and without melody, it is essential to standardize these elements across participants. To achieve this, a musical piece was created that incorporates all of the above considerations.

The tempo will be determined during the pre-intervention phase for each participant using a baseline 10-meter walk assessment and will be applied during the intervention phase. Using online composing software Noteflight, we will dynamically set the beats per minute (bpm) of pre-composed music to match their normal gait tempo and repeat with a 10% faster metronome beat. The remaining elements of the composition will include melody, pitch, timbre, and rhythm. This approach will allow for precise adaptation of musical elements while maintaining the core structure of the composition (see Supplementary material 1).

For the melodic RAS, the composed music incorporated key musical elements that align with therapeutic goals, drawing on findings from previous research (Creel, 2020; Štillová et al., 2021; Weiland et al., 2011). Table 2 outlines the primary components of the composed melodic RAS music.

**Table 2.** Key Components of Melodic RAS music.

| Key components of composed music for Melodic RAS condition |
| --- |
| • Use a 2/2 meter to mimic a march<br>• Over 50% beat correlation with an idealized 2/2 meter<br>• Avoid dissonance<br>• Clear melody<br>• Emphasis on harmony<br>• A clear contour with a balance of rising and falling phrasing and sense of development<br>• Additional percussion part to keep the pulse clear according to the RAS BPM (Beats per Minute) determined by the pre-intervention phase |

### Study protocol

This study protocol has been registered on ClinicalTrials.gov (Identifier: NCT05763732). After participants provide consent, they will complete the Barcelona Music Reward Questionnaire (Mas-Herrero et al., 2012) to assess their perceived level of reward experienced with music, helping us understand how individual music preferences and emotional connections to music might influence RAS intervention outcomes. The protocol consists of two conditions (DBS ON and DBS OFF), each of which is divided into a Pre-RAS phase, a RAS/Intervention phase and a Post-RAS phase. The order of stimulation states will be block randomized among participants.

Figure 1 illustrates the study protocol. LFP recordings will be collected across all stages. Each bracket in Figure 1 is considered an “event” and is subsequently saved as a .json file. Consequently, the protocol encompasses 26 events, including 1) pre initial walk, 2) initial walk, 3) post initial walk, 4) pure RAS walk, 5) rest, 6) melodic RAS walk, 7) rest, 8) pure-RAS 110% tempo walk, 9) rest, 10) melodic-RAS 110% tempo walk, 11) pre final walk, 12) final walk, 13) post final walk; 14) - 26) repeat 1) – 13) for either DBS ON or OFF). In the DBS ON condition, participants will receive optimized stimulation unless the active contact is at a lead end, preventing brain sensing. In such cases, parameters are modified to permit sensing electrode placement above and below stimulation, while closely mirroring the original optimized settings. After an initial rest period of 2 minutes during which the patient will be sitting in a chair, the study clinician performs the MDS-UPDRS-III rating scale part 3.9 through 3.13. Following this assessment, participants will begin a series of 2-minute walks with breaks in between.

**Figure 1.**
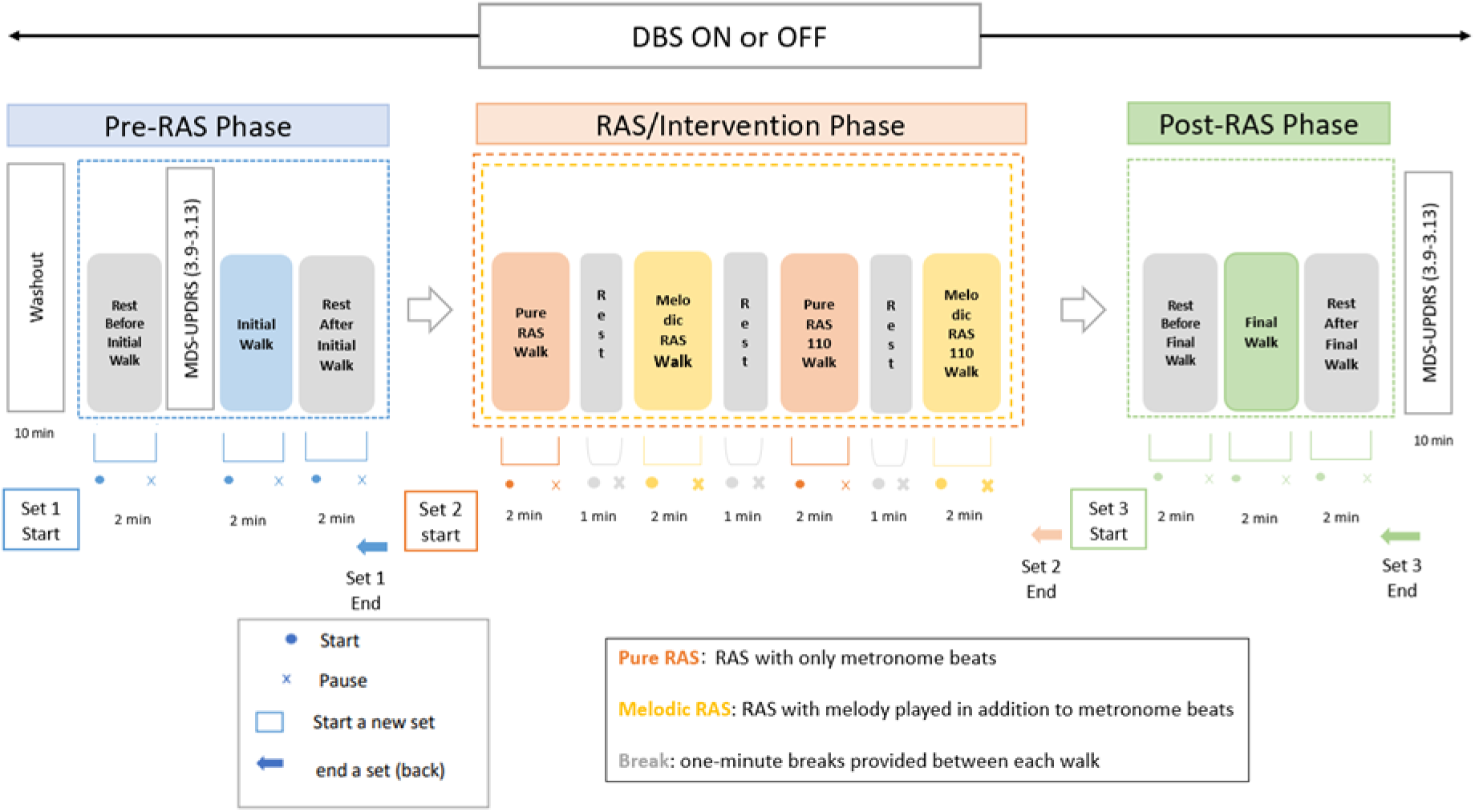
**Study Protocol**

The first walk, titled “Initial Walk” in Figure 1, will be used to set the baseline cadence, where participants will be asked to walk for 2 minutes on a set path without any intervention. The cadence measured during this walk will be used to determine the tempo at which the subsequent RAS interventions are provided. Measuring the baseline cadence for each participant in this fashion will allow for individual adjustments for each participant, rather than setting a common tempo for all participants (Calabrò et al., 2019). When comparing the different RAS interventions, the LFP measured during this initial walk will serve as the baseline measurement. This reference point will allow for comparison of neural activity under different conditions, facilitating an understanding of how each type of RAS influences subthalamic or pallidal LFP activity and associated motor performance.

After another 2-minute resting period, participants will enter the intervention phase. The first half consists of walking to metronome beats set to the base cadence. This includes the pure RAS intervention, where participants will walk to metronome beats (“Pure RAS”) and the melodic RAS intervention, where participants will listen to an original composition at the same tempo with superimposed metronome beats while walking. The same task will be repeated, but with a 10% increase in tempo relative to the base cadence, following a standard RAS protocol (M. Thaut, 2013; M. Thaut & Hoemberg, 2014). To account for the sequence of having either the pure or melodic RAS walk first, the order in which the participants first walk with melody or without melody will be randomized.

Finally, during the post-RAS phase, participants will undergo the same procedures as in the pre-RAS phase. By comparing the outcomes between the pre-RAS and post-RAS phases, we aim to determine whether any immediate effects of the interventions persist beyond their immediate application.

In DBS OFF, there will be a 10-minute washout period to allow the participant’s brain circuits to adjust to the absence of stimulation. Aside from the stimulation state, DBS OFF phase will follow the same protocol as the DBS ON phase.

After completing the interventions, participants will be asked to fill out a brief post-session survey to assess the subjective impact and perceived differences between the pure RAS and melodic RAS interventions (Supplementary material 2).

### Data Collection

#### Demographic Data

Participants’ demographic data will be obtained from their electronic medical record. Key variables include age, gender, years since PD diagnosis, medication status, asymmetry of symptoms if relevant, DBS target and years since DBS surgery. Additionally, participants will be asked to report the time of their last medication dose on the day of data collection.

#### Behavioral Data

During the 10-meter walk assessment over 2 minutes, cadence and velocity will be derived by averaging the results of the three 10-meter trials, focusing on the middle section of the 2-minute walk. This method aims to minimize variability and provide more reliable gait measures.

The study clinician will assess MDS UPDRS part III items 3.9 through 3.13 directly before the initial walk in the pre-RAS phase and directly after the rest after final walk in the post-RAS phase (Figure 1). This will be repeated during DBS ON and DBS OFF conditions.

#### LFP Data

The Medtronic Percept™ PC allows for the measurement of LFP through DBS leads implanted in the brain, even when the DBS is turned OFF. There are four electrodes on each lead in each hemisphere (left: 0 to 3; right: 8 to 11). To use BrainSense^TM^ Streaming technology and record LFP, the electrode configuration must be compatible. During sensing, only the middle two contacts (left: 1 or 2 and right: 9 or 10) can be used for actual stimulation (Feldmann et al., 2022), and the recording electrodes must be set to above and below the active electrodes (left: 0 and 3 or right 8 and 11) (Figure 2).

**Figure 2.**
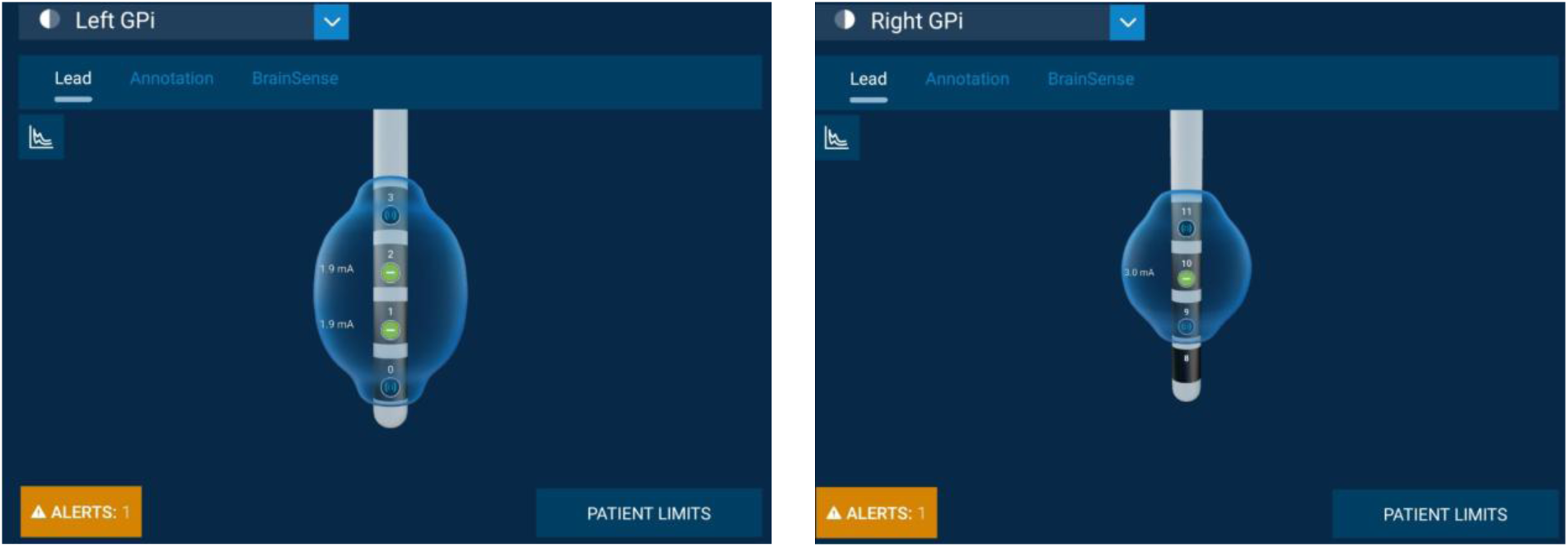
**Example of electrode setup for stimulation and LFP recording in GPi**.

If the patient’s clinical settings utilize electrodes 0,3,8, or 11 for stimulation electrodes, the BrainSense Group configuration must be adjusted so that the simulating contact is one of the two middle contacts. This configuration will be used during the entirety of the study session. Following the session, participants will be returned to their original clinical DBS settings. The setup process will begin with an impedance test, followed by a signal test during which the LFP is read. The clinician then will select the contact pair with the largest peak in the beta frequency range, which will determine a frequency band of interest (5Hz range of the selected frequency). The power of this frequency band across time will be recorded at a sample rate of 2Hz and provided in the output as BrainSense LFP. During the study visit itself, streaming technology will be used to collect time domain data and to observe changes in LFP in response to different activities.

Another advantage of the streaming function is that it does not impose a strict limit on recording duration. However, to reduce the risk of potential issues—such as longer processing times for JSON exports or possible data loss—we will record in 10-minute segments for each session. Thus, new sets will be made for each of the different phases: Pre-RAS Phase, Intervention Phase, and Post-RAS Phase. In this setting, the LFP data will be streamed from both hemispheres, and the time domain data will be sampled at a rate of 250Hz. After the end of the session, the “Export Json Session Data” will be selected. The .json file report can subsequently be opened and analyzed using MATLAB.

### Data Analysis

Both behavioral measures and LFP activity will be compared across the *walking* events: initial walk (no RAS), during pure RAS (at base cadence and at 110% base cadence), during melodic RAS (at base cadence and at 110% base cadence), and final walk (no RAS) in DBS ON and DBS OFF state.

#### Behavioral Data

The number of steps and the amount of time a participant needs to walk 10 meters (two rounds of 5 meters) will be used to calculate the velocity and the cadence (cadence=(60/seconds) *(number of steps)). For each walking trial, the participant’s cadence will be calculated as the average of three recordings taken from the middle segment of the walking bout, starting at 40 seconds. This approach is intended to capture cadence data that reflects steady-state walking, rather than the initial phase. Rhythmic entrainment (how well a listener is able to match a given tempo) will be measured by the percentage change divergence between the participant’s measured cadence and the given tempo during their intervention phase (Figure 3A). The lower the absolute value of this percentage change, the better entrained the participant is to the given tempo.

**Figure 3.**
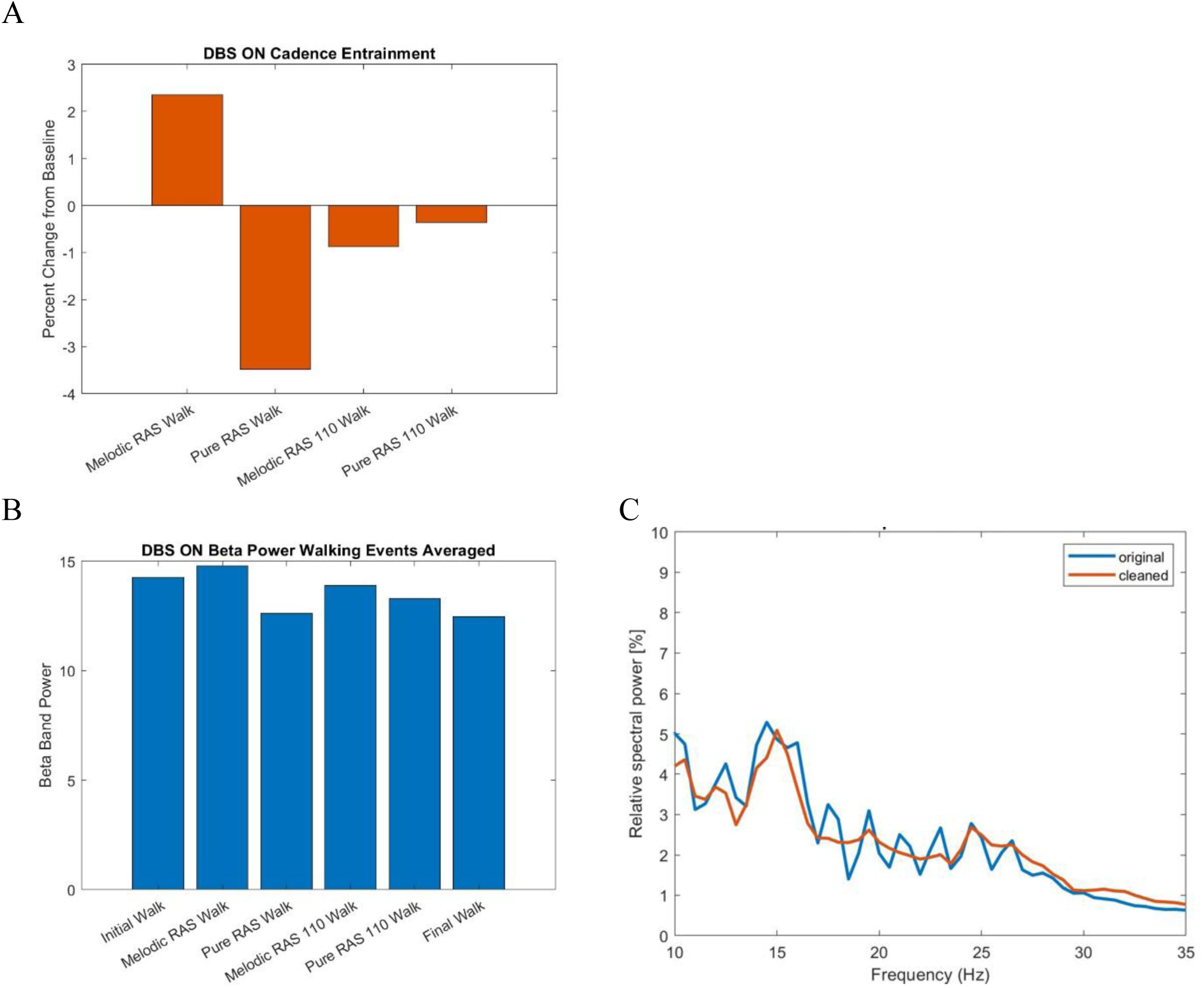
**Representative examples of: A. Changes in behavioral entrainment levels; B. Beta band power; C. Power spectral density.**

#### LFP Data

There is currently no single industry standard for LFP analysis, nor an established measurement for studying the effect of RAS on LFP during walking. Additionally, while suppression of beta power has been shown in multiple studies to be an increasingly accepted biomarker for improvement in PD, this has more commonly been shown for symptoms such as bradykinesia (Feldmann et al., 2022) and rigidity (Kühn et al., 2009), rather than gait. One paper by Storzer et al. reported on the potential relevance of a small band around 18Hz that could be relevant for freezing of gait (Storzer et al., 2017). However, there is currently not enough literature to warrant determining the severity of gait impairment by purely using LFP. Thus, this study will employ various methods of LFP analysis to identify the outcome measure that most accurately reflects the effects of RAS. The goal is to establish an LFP metric that could be used as a biomarker for assessing gait improvement in future research. All LFP analysis will be performed in MATLAB.

Similar to other neurophysiological measures, LFPs are susceptible to potential artifacts. In the case of LFP, common sources of such artifacts are cardiac activity and movement artifacts (Neumann et al., 2021). There are different methods to address these artifacts. In this study, we will be using the open source Perceive toolbox (https://github.com/neuromodulation/perceive) (Neumann et al., 2021) to both correct for artifacts and support other parts of the LFP analysis. Although no existing tool or subject matter expert can fully eliminate all artifacts, this toolbox has demonstrated efficacy in reducing artifact-induced power, thereby enhancing the analysis of LFP dynamics (Stam et al., 2023)

Initially, our focus will be on beta power (Doyle et al., 2005; Kühn et al., 2006b) (Figure 3B). Existing literature also indicates that subbands within the LFP spectrum may be more closely linked to symptom improvement, with some studies suggesting that specific subbands are associated with particular PD symptoms. Therefore, we will examine the effect of various RAS interventions on the overall LFP beta power (13 to 35Hz), as well as on the low beta band (13-20Hz) and high beta band (21-35Hz).

Another outcome measurement used in previous studies is the peak approach. Peaks are generally defined as the “maxima in normalized spectral power” (Giannini et al., 2023). However, the method to determine peaks has differed across studies. Typically, when focusing on peaks, the power spectrum density (PSD) is used to find peaks either visually (Feldmann et al., 2022) or through an original algorithm (Hell et al., 2018). In comparing PSD across patients, analyzing relative power was suggested over absolute power, as the latter can be more susceptible to local signal variability and external influences (Neumann et al., 2016) and will thus be inherently different for each patient according to the placement of their leads during DBS surgery. Therefore, we will first produce PSD figures in the beta band (13 to 35 Hz) for each intervention type using the Perceive toolbox, which will then be visually inspected for the presence of peaks (Figure 3C). When such a peak is found, the power at +/-2.5Hz of that frequency will be calculated (Özkurt et al., 2011).

While the beta band has been widely used in LFP analysis, the gamma band has received less focus. Some studies report that the gamma range is associated with a prokinetic effect in PD (Florin et al., 2013; Swann et al., 2016). Therefore, we will also investigate if there is a relevance of RAS on the low gamma band, limited to the 31 to 95 Hz range. This limitation is due to the 250 Hz sampling frequency of the time domain data collection of the Percept™ PC or RC device.

#### Statistical Analysis

The statistical analysis will be performed in four different stages using the software R. Our statistical analysis plan has been shaped by specific limitations, which include small sample size, repeated measures, and the likely nonparametric distribution of collected data. To address these factors, we will proceed with the following statistical analysis plan: 1) Assess the effect of different RAS interventions (initial walk, pure RAS, melodic RAS, pure RAS 110, melodic RAS 110, final walk) on LFP measurements using the Friedman Test; 2) Evaluate the impact of various RAS interventions on gait parameters, specifically cadence entrainment and velocity, also using the Friedman Test; 3) Investigate the correlations between LFP measurements and gait parameters employing a Linear Mixed Model (LMM). The demographic information gathered (age, sex, disease duration) will be taken into consideration in the LMM as random effects.

Additionally, as an exploratory analysis, we will examine whether RAS with melody or RAS-first interventions result in differing effects on gait parameters and LFP changes by including the order of types of RAS as a fixed covariate. We acknowledge that the small sample size may limit the statistical power of LMM analysis. To address this limitation, we will conduct a sensitivity analysis, including comparisons between simplified and full model specifications, leave-one-out analyses to examine the influence of individual participants, and bootstrap resampling to assess the stability of parameter estimates. These steps will help evaluate the robustness of our findings despite the constraints of the sample size.

#### Data management plans

All study data will be collected and stored in compliance with institutional and federal data protection regulations, including HIPAA. Behavioral, neurophysiological (LFP), and clinical assessment data will be de-identified and coded using a secure participant ID system. Data will be entered into and managed through a secured Johns Hopkins OneDrive server.

LFP data captured during triggered events will be stored as the implantable pulse generator (IPG) and downloaded to the clinician programmer. Data will be then exported as .json files, which can be imported into MATLAB for preprocessing and analysis. The files will be subsequently stored in a secure location such as JH OneDrive, accessible only to IRB-approved study personnel.

All digital data files contain only numerical behavioral performance results and no identifying information. Any additional digital files, including raw or processed neurophysiological data, will be housed and analyzed within a dedicated JH OneDrive folder shared exclusively with the approved study team. Access to these storage locations is restricted and must be granted by the Principal Investigator. All data encryption and security protocols meet or exceed HIPAA compliance standards. Data will not be shared outside of Johns Hopkins University. Regular data audits will be conducted to ensure accuracy, completeness, and compliance with the approved protocol.

#### Safety considerations

Given the involvement of individuals with implanted DBS systems, safety protocols have been established in close consultation with neurology, neurosurgery, and neurologic music therapy collaborators. RAS stimuli will be carefully calibrated to avoid excessive auditory stimulation or entrainment effects that could interfere with motor function or induce discomfort.

No changes will be made to the participants’ clinical DBS settings, except in cases where the top or bottom contact is used clinically and one of the middle contact levels will need to be used to allow LFP recording or streaming; in these cases, a temporarily stimulation profile will be made uniquely for the study session, and they will be switched back to their original stimulation profile after the study session. All sessions will be conducted under clinical supervision, and participants will be monitored for any adverse effects, including fatigue, dizziness, or changes in mood or motor symptoms during the session.

#### The status and timeline of the study

This study is currently in the preparatory phase, with preliminary data collected from seven participants with STN DBS implants with Percept PC. These initial recordings have informed feasibility and protocol refinement. Data analysis pipelines for advanced LFP processing are currently under development, including custom scripts for signal extraction and event-related analyses. Full data collection is anticipated to continue over a 12-month period following recruitment, with subsequent phases dedicated to data cleaning, statistical analysis, and dissemination of findings. Preliminary results may be shared at relevant scientific conferences, and the full study outcomes will be submitted for peer-reviewed publication upon completion.

## Discussion

This study will aim to investigate the impact of pure- and melodic-RAS during DBS ON and OFF on patients with PD who have the Medtronic Percept^TM^ PC or RC DBS device with brain leads in the STN or GPi. The main innovation of this study lies in the use of LFP to deepen our understanding of the neurophysiological effects of RAS in PD patients with DBS. Additionally, the Medtronic Percept^TM^ PC or RC DBS device allows LFP to be recorded during the DBS OFF stage, which is an important and novel addition in comparison to other existing DBS devices. Finally, the use of an original composition for RAS with the flexibility to change the tempo in the context of RAS is also novel, allowing the researchers to focus on the effect of melody on RAS while keeping other factors in a musical piece (such as familiarity, timber and rhythmic structure) constant.

The goal of the study is to recruit 10 participants each with STN and GPi DBS. This number of participants is considered feasible for the following reasons: First, there are many DBS devices available on the US market, including Boston Scientific and Abbott. The Percept device was first developed in 2020, and many patients still use its precursor, the Activa PC. Second, the implantation of the DBS device is commonly split among the STN and the GPi to address specific predominant symptoms of PD. Third, for some patients with DBS, their degree of gait impairment has progressed to a point where they no longer fulfill the inclusion criteria for the study. These difficulties have been experienced during the recruitment process. We will aim to recruit 20 participants for the purposes of this pilot study. Despite this limitation, the effect size and sample size estimates derived from this pilot study can provide valuable insights for future investigations into RAS. In addition, the preliminary data will allow us to refine our understanding of both the neurophysiological and behavioral responses to RAS, thereby informing us of the design and scope of future studies. More importantly, by examining the relationship between LFP activity and gait parameters across different RAS conditions (e.g., pure tone vs. melodic cues), this study aims to identify the types of auditory stimulation that elicit the most adaptive neural responses—such as suppression of pathological beta oscillations or enhancement of movement-related gamma activity. These findings may contribute to the development of individualized cueing strategies based on patients’ neural profiles, potentially optimizing therapeutic outcomes. Furthermore, this line of research could inform the design of closed-loop DBS or neurofeedback systems that dynamically adjust auditory cues or stimulation parameters in response to real-time LFP signals during walking. Such approaches may improve gait performance, reduce freezing of gait, and facilitate the translation of RAS interventions from controlled clinical settings to everyday mobility contexts.

### Dissemination Plans

The results of this study will be disseminated through peer-reviewed publications in relevant neuroscience, rehabilitation, and music-based intervention journals. Findings will also be presented at national and international scientific conferences, including those focused on neuromodulation, auditory neuroscience, and music therapy. To promote transparency and reproducibility, de-identified datasets, analysis scripts, and relevant materials (e.g., auditory stimuli, software interface screenshots) will be shared via open-access platforms such as the Open Science Framework (OSF), contingent upon ethical approval and participant confidentiality considerations. In addition, lay summaries will be provided to participants and community stakeholders when appropriate.

### Protocol Amendments and Study Termination

Any modifications to the study protocol, including changes in study design, eligibility criteria, outcome measures, or data collection procedures, will be documented and submitted for prior approval by the Institutional Review Board (IRB). Substantial amendments will be clearly indicated in future publications and version-tracked using protocol registries, where applicable. In the event of study termination—whether due to safety concerns, logistical challenges, or lack of feasibility—all data collected to that point will be securely archived and analyzed in accordance with the original study aims, unless otherwise restricted. The reasons for termination will be transparently reported in the final manuscript and any public data repositories, as appropriate.

## Declarations

The authors declare that there are no disclosures to report.

## Funding Statement

No external funding was received for this study.

## Data Availability

All data produced in the present study are available upon reasonable request to the authors.

## Notes

### Competing Interest Statement

The authors have declared no competing interest.

### Clinical Trial

NCT05763732

### Funding Statement

This study did not receive any funding.

### Author Declarations

Ethics committee/IRB of Johns Hopkins University School of Medicine gave ethical approval for this work.

