## Supplementary figures and images for "Effect of Rhythmic Auditory Stimulation (RAS)^®^ with and without Melody on Parkinson’s Disease (PD) Patients with Deep Brain Stimulation (DBS): A Study Protocol"

### Supplementary Material 1

#### Happy March version

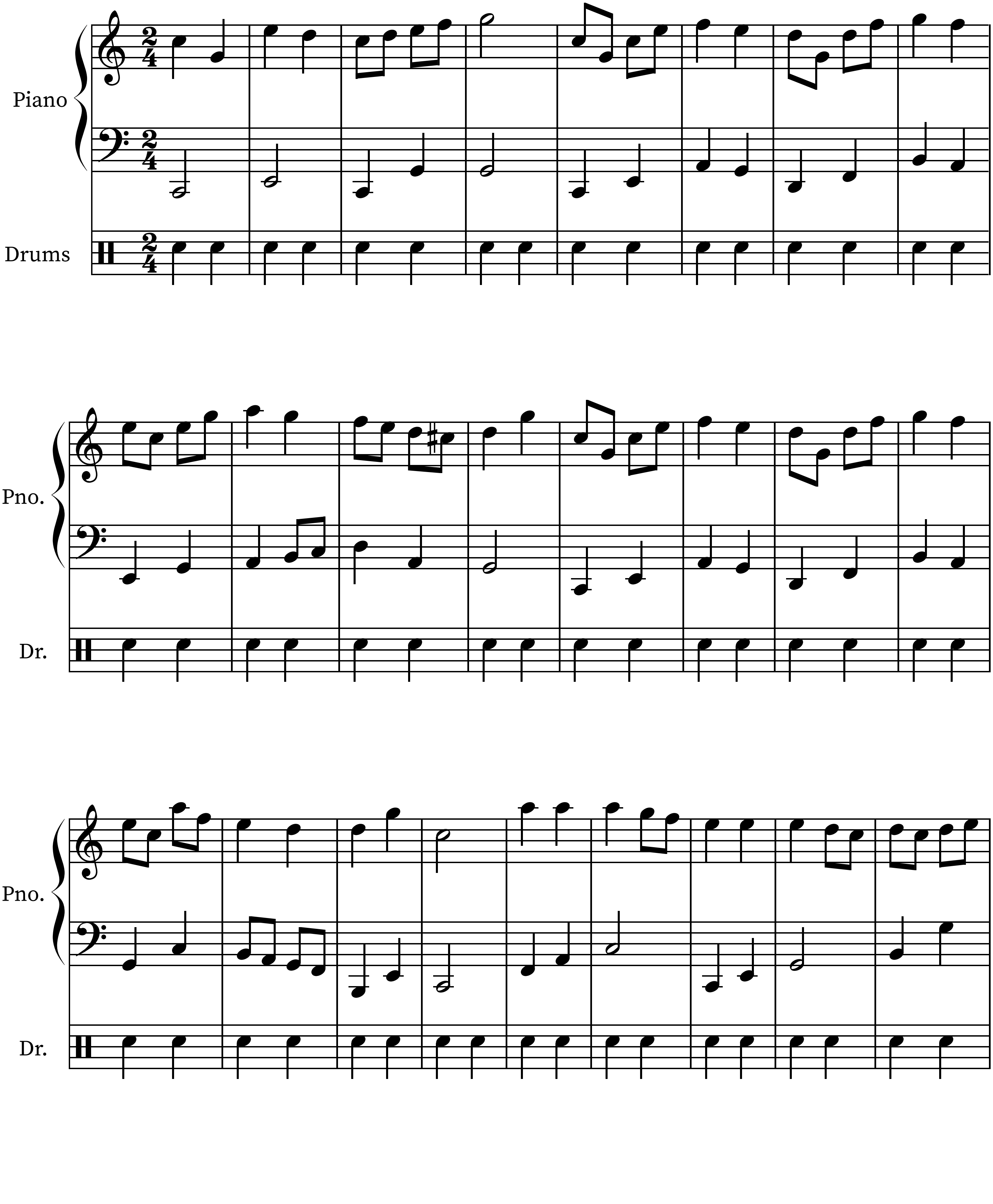

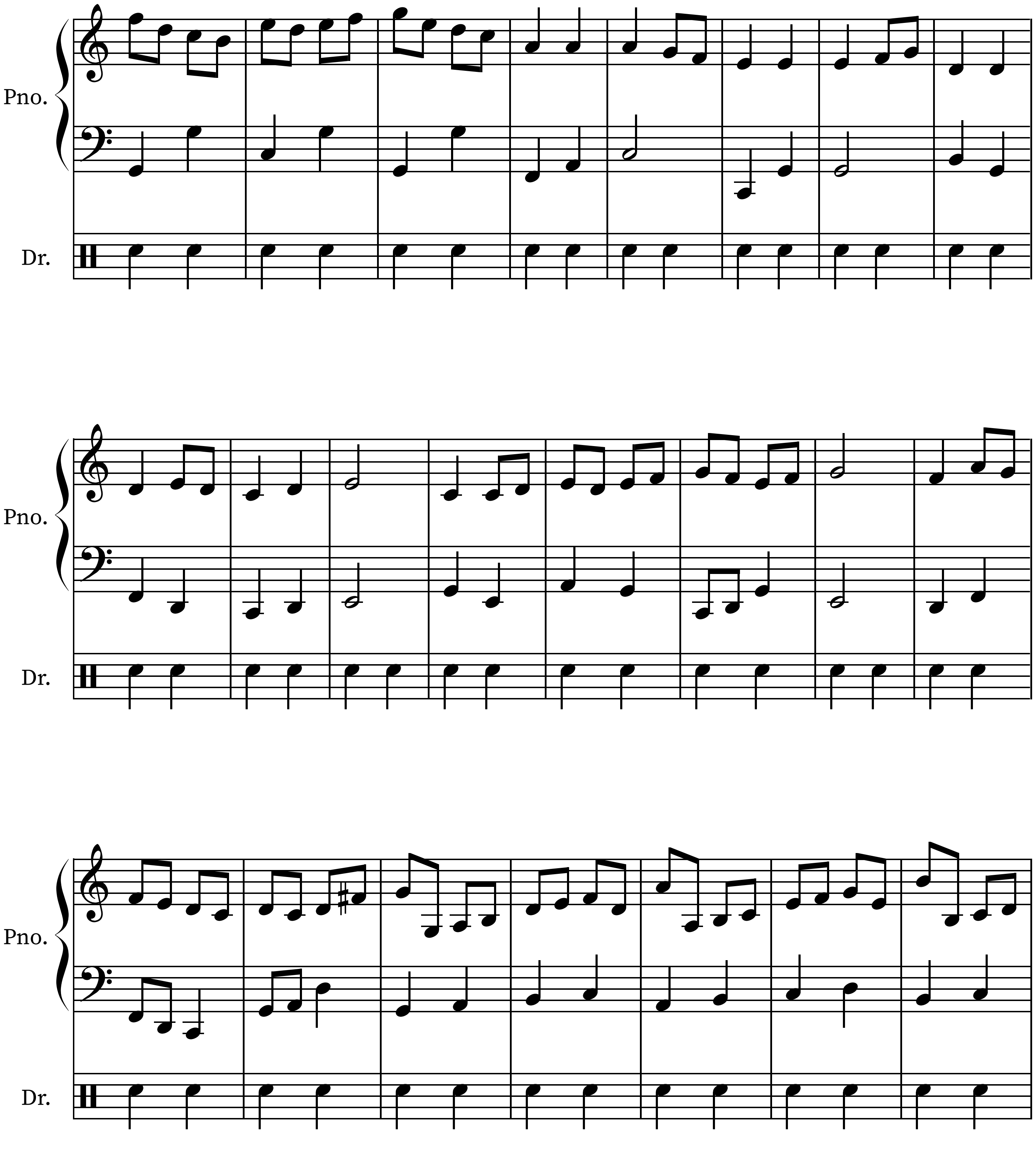

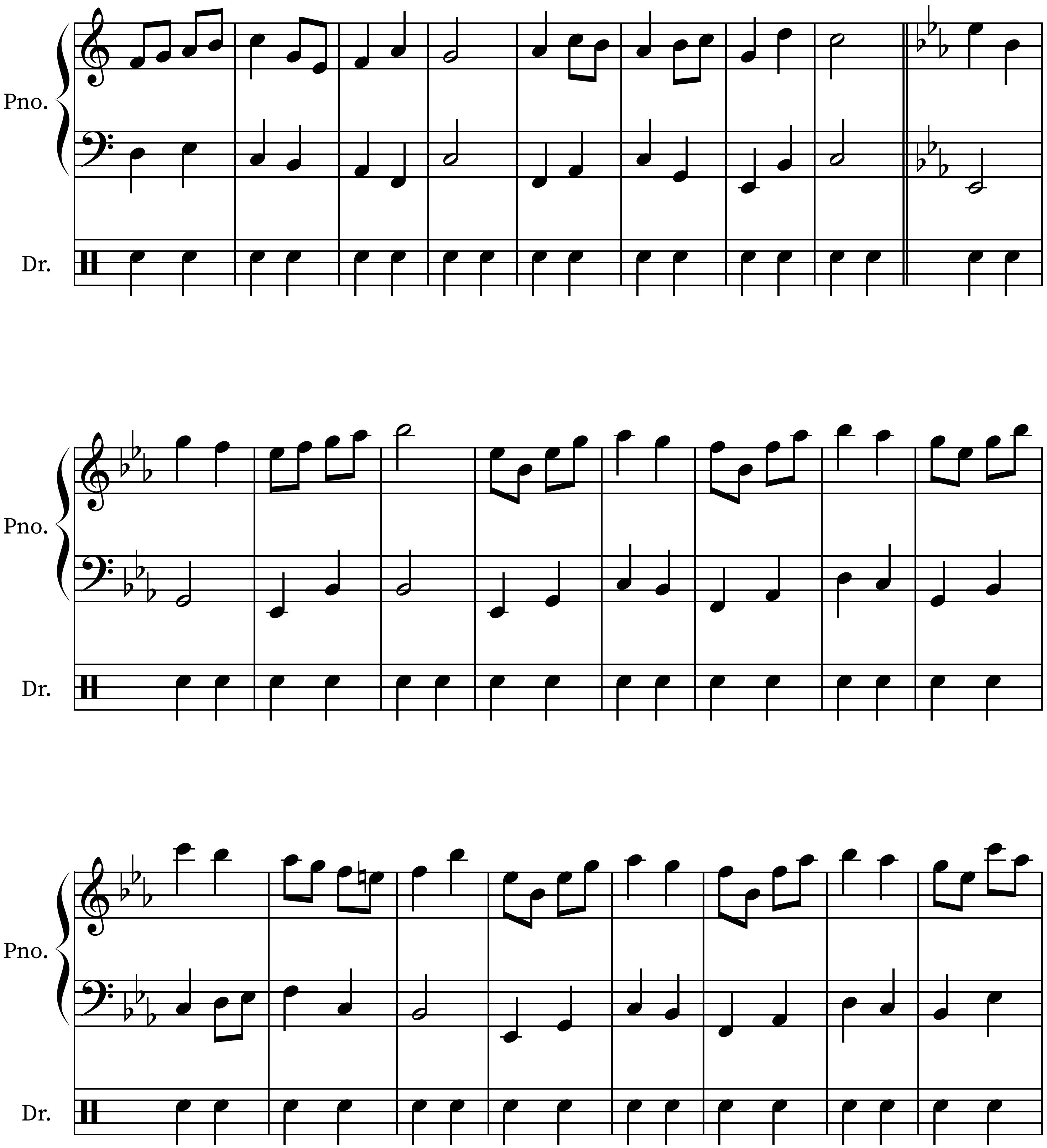

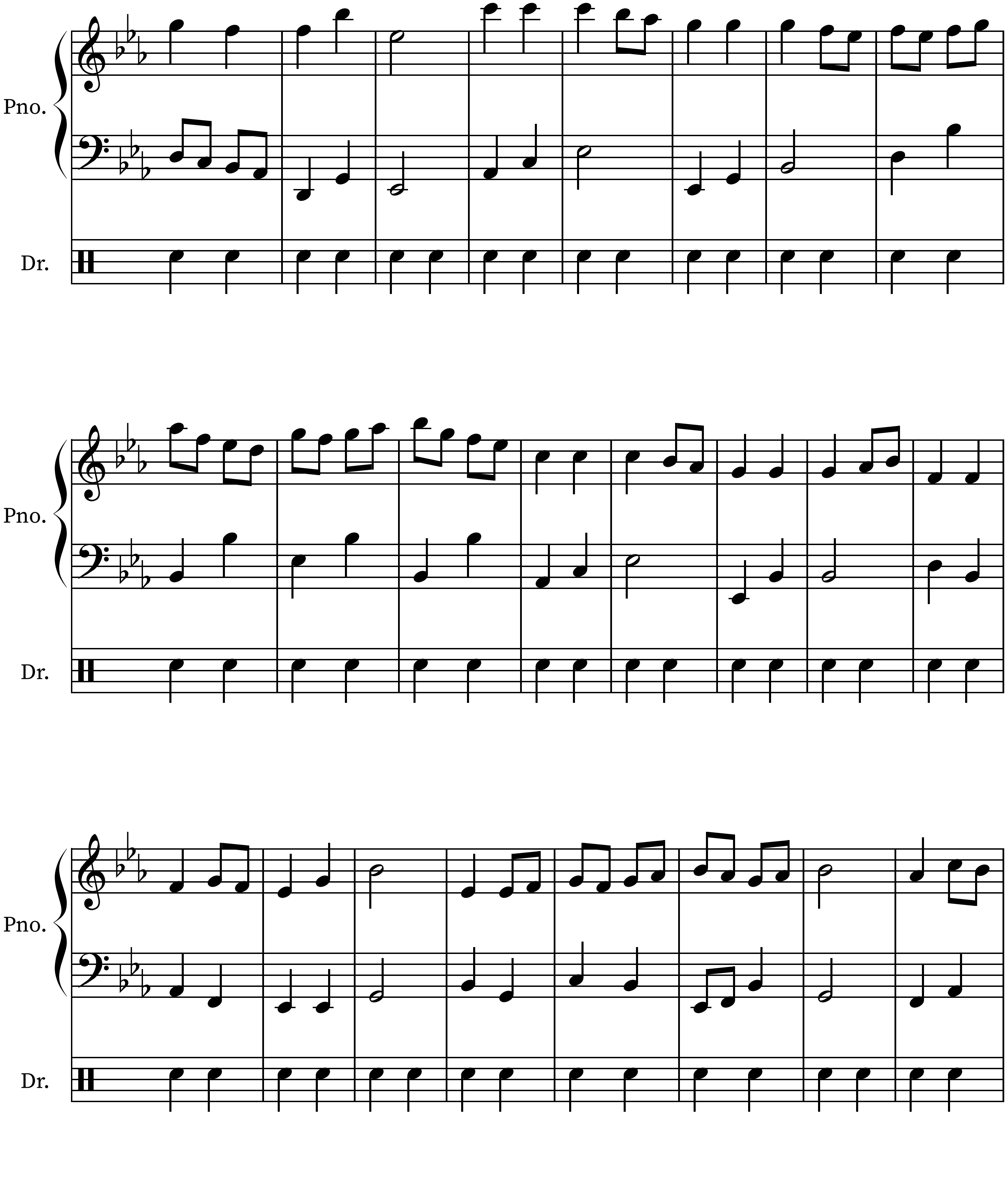

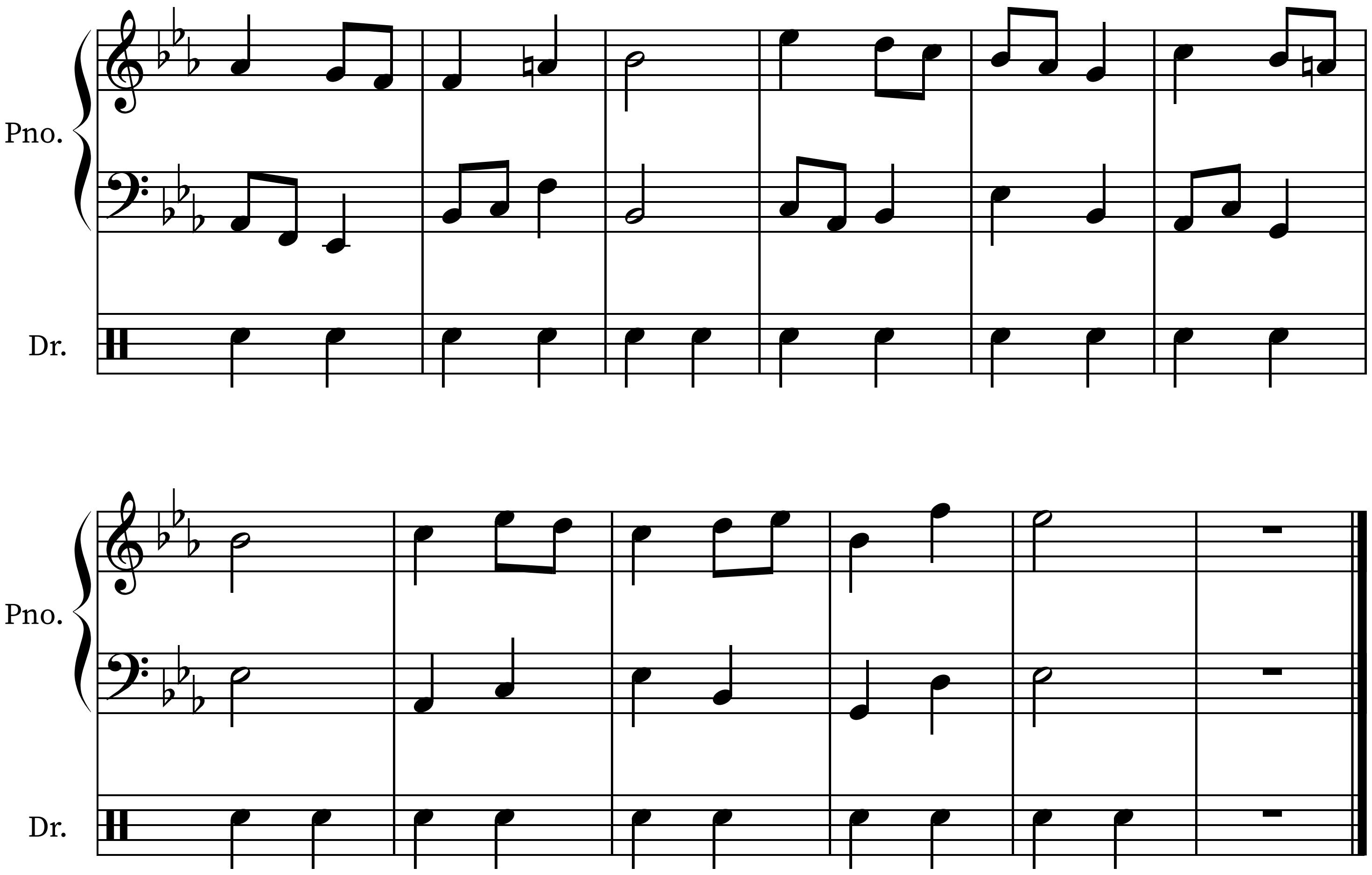
