## Supplementary Material 2 for "Effect of Rhythmic Auditory Stimulation (RAS)^®^ with and without Melody on Parkinson’s Disease (PD) Patients with Deep Brain Stimulation (DBS): A Study Protocol"

Survey Questions

Part 1: open ended questions

1. Please describe any specific feelings or thoughts you had while walking to rhythmic beats with and without melody.

2. Did the melody remind you of anything?

3. How did the rhythmic beats with and without melody affect your walking experience, if at all?

4. Were there any challenges you faced while walking to either rhythmic beats with or without melody? Please explain.

Part 2: scaled questions

Please answer the following questions on a scale of 1 to 5

1 strongly disagree

2 disagree

3 neutral

4 agree

5 strongly agree

1. The rhythmic beats **without** melody were easy to follow.

   1 – 2 – 3 – 4 – 5

2. The melody **with** rhythmic beats was easy to follow.

   1 – 2 – 3 – 4 – 5

3. The rhythmic beats **without** melody helped me walk less rigidly/stiffly.

   1 – 2 – 3 – 4 – 5

4. The melody **with** rhythmic beats helped me walk less rigidly/stiffly.

   1 – 2 – 3 – 4 – 5

5. I would like to use rhythmic beats **without** melody at home.

1 – 2 – 3 – 4 – 5

6. I would like to use the melody **with** rhythmic beats at home.

1 – 2 – 3 – 4 – 5

7. The rhythmic beats **without** melody helped take my mind off of the task at hand (walking).

1 – 2 – 3 – 4 – 5

8. The melody **with** rhythmic beats helped take my mind off of the task at hand (walking).

1 – 2 – 3 – 4 – 5

9. I was more confident walking with the rhythmic beats **without** melody.

1 – 2 – 3 – 4 – 5

10. I was more confident walking with the melody **with** rhythmic beats.

1 – 2 – 3 – 4 – 5

11. I was able to track the beat better with the melody as opposed to without the melody.

1 – 2 – 3 – 4 – 5

12. Is there anything else you would like to share about your experience in this study?
